# Testing & Opening in Augustusburg A Success Story?

**DOI:** 10.1101/2021.06.16.21258869

**Authors:** Marc Diederichs, Timo Mitze, Felix Schulz, Klaus Wälde

**Affiliations:** Johannes Gutenberg University Mainz; University of Southern Denmark; CESifo and IZA

## Abstract

The city of Augustusburg allowed for opening of, inter alia, restaurants and hotels joint with large-scale testing. We evaluate this testing & opening (T&O) experiment by comparing the evolution of case rates in Augustusburg with the evolution in other communities of Saxony. We have access to small-scale SARS-CoV-2 infection data at the community level (” Gemeinde”) instead of the county level (” Landkreis”) usually used for disease surveillance. Despite data challenges, we conclude that T&O did not lead to any increase in case rates in Augustusburg compared to its control county. When we measure the effect of T&O on cumulative cases, we find a small increase in Augustusburg. This difference almost completely disappears when we control for the effect of higher case rates due to more testing. Generally speaking, T&O worked much better than in comparable projects elsewhere.

---

Augustusburg is a town in Saxony with approx. 4500 inhabitants. It implemented an innovative IT-supported public-health project as of April 1, 2021 consisting of large-scale testing joint with opening of, inter alia, restaurants and hotels. We evaluate this testing & opening (T&O) experiment in two steps. First, we compare the evolution of the pandemic in Augustusburg with its evolution in other communities of Saxony. Second, we look at individual data of visitors and inhabitants of Augustusburg. The latter comprise data on testing (with positive and negative results) and data on visits of restaurant, hotels and other participating institutions.

The data is in principle remarkable in two senses. First, we have access to small-scale SARS-CoV-2 infection data at the community level (” Gemeinde”) instead of the county level (” Landkreis”) usually used for disease surveillance. Second, individual data was collected by local authorities in Augustusburg (supported by IT firm Theed). We observe visitors and inhabitants participating in the project by the minute during the day.

In practice, data provides some challenges. When we compare case rates across communities in Saxony, we find that data quality issues at the community level are more visible than at the county level. This is true for reported seven-day case rates and for computed daily cases.

Accepting potential measurement error in the data, we find that T&O did not lead to any increase in case rates in Augustusburg compared to its control county. When we measure the effect of T&O on cumulative cases, we find a small increase in Augustusburg. This difference almost completely disappears when we control for the effect of higher case rates due to more testing. The difference in cumulative cases between Augustusburg and its control county is not statistically significant.

Generally speaking, T&O worked much better than in comparable projects elsewhere. This might be due to the design of T&O in Augustusburg: First, participants did not only have to present a negative test before participating in a T&O event but also had to register for every event. This makes sure that only negatively tested individuals participate in events. Second, participants were tested every day. The implied identification of positive cases introduced another layer of safety.

In contrast to other experiments, Augustusburg collected a wealth of individual data as described above. These data are subject to ongoing research and results will be made available as soon as possible. T&O ended on Saturday April 24 due to the ‘Bundesinfektionsschutzgesetz’ adopted by the German federal parliament.

## 1 Findings

### 1.1 Data quality

One of the promising features of this research project consisted in the usage of data at the community rather than at the county level. As communities are much smaller units than counties (Saxony consists of 13 counties but 419 communities), the effect of a public health experiment should be measurable either much more easily or with higher precision. The presence of many communities under almost identical policy rules of one federal state should also facilitate the identification of control regions compared to other projects where data is available only at the county level. On the other hand, data at the community level must be reliable. One of our results therefore concerns data quality. We distinguish between daily seven-day case rates and daily cases.

- Daily seven-day case rates Data at the community level is provided online [8] by the Saxon State Ministry for Social Affairs and Social Cohesion (‘Sächsisches Staatsministerium für Soziales und Gesellschaftlichen Zusammenhalt’). Users are encouraged to use data with care. When inspecting this data e.g. for Augustusburg (see appendix A.1.2), we see that seven-day case rates contain spikes which, by construction of case rates, cannot occur. As measurement problems exist in all data sets, we assume that these are minor consistency issues and start from these community-level case rates for our first analysis of the effect of T&O.
- Daily cases As daily cases were not available, we computed daily cases from seven-day case rates (see appendix A.4.2 for the theoretical background). This analysis showed that negative daily cases easily result from our procedure. While we might always be wrong with our procedure, we do conclude that this is a further indication that data needs to be used with care. All of our findings are therefore conditional on data quality.

### 1.2 The effects of testing and opening based on case rates

Does the opening of restaurants and hotels joint with regular rapid testing as of April 1 in Augustusburg lead to an increase in the number of SARS-CoV-2 infections? We answer this question by comparing the case rate of Augustusburg, as it actually evolved under T&O, with the case rate of Augustusburg as it would have evolved without T&O. Since this counterfactual situation of Augustusburg is not observable, we employ the ‘synthetic control method’ (SCM) explained in more detail in section 2.

Figure 1 shows the effect of T&O on daily case rates (see appendix A.4.2 for a formal definition of case rates and related pandemic measures). The figure compares the case rate in Augustusburg before and after April 1 with a ‘synthetic twin community’ of Augustusburg. SCM has identified a relatively large number of (ten) control communities, shown in table 1. The selection of control communities is based on criteria (‘predictor set’) shown in table 4. Looking at the respective weights of control communities shows that one community dominates with a weight of 33% while the remaining 9 communities have similar weights between 11% and 4%. These 10 communities form the synthetic twin of Augustusburg.

**Table 1:** Weights for figure 1

| Name | Weight |
| --- | --- |
| Regis-Breitingen, Stadt | 0.33 |
| Weißenberg, Stadt | 0.11 |
| Thalheim/Erzgeb., Stadt | 0.094 |
| Sayda | 0.094 |
| Eilenburg-West | 0.085 |
| Brand-Erbisdorf, Stadt | 0.073 |
| Leutersdorf | 0.067 |
| Hoyerswerda, Stadt | 0.055 |
| Wechselburg | 0.044 |
| Bad Elster, Stadt | 0.042 |

**Table 2:**
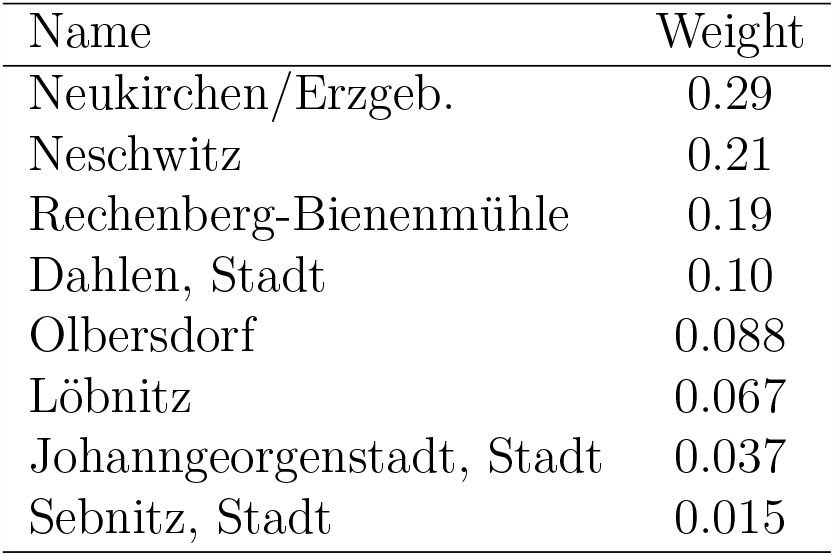
Weights for figure 2

**Table 3:** Positive tests 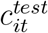, test rate 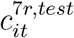, correction of case rate 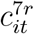 and total number of tests 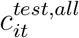

| date | $c_{it}^{test}$ | $c_{it}^{7r,test}$ | $c_{it}^{7r}$ | $\hat{c}_{it}^{7r}(50\%)$ | $\hat{c}_{it}^{7r}(80\%)$ | $c_{it}^{test,all}$ |
| --- | --- | --- | --- | --- | --- | --- |
| Apr 1 | 1 | 0 | 332 | 332 | 332 | 3 |
| Apr 2 | 0 | 22 | 332 | 321 | 315 | 0 |
| Apr 3 | 0 | 22 | 332 | 321 | 315 | 1 |
| Apr 4 | 0 | 22 | 332 | 321 | 315 | 1 |
| Apr 5 | 1 | 22 | 332 | 321 | 315 | 1 |
| Apr 6 | 1 | 44 | 332 | 310 | 297 | 6 |
| Apr 7 | 0 | 66 | 332 | 299 | 279 | 10 |
| Apr 8 | 0 | 66 | 310 | 277 | 257 | 9 |
| Apr 9 | 0 | 44 | 355 | 332 | 319 | 18 |
| Apr 10 | 0 | 44 | 421 | 399 | 386 | 7 |
| Apr 11 | 0 | 44 | 332 | 310 | 297 | 1 |
| Apr 12 | 3 | 44 | 332 | 310 | 297 | 18 |
| Apr 13 | 0 | 89 | 332 | 288 | 262 | 6 |
| Apr 14 | 0 | 66 | 421 | 388 | 368 | 10 |
| Apr 15 | 0 | 66 | 421 | 388 | 368 | 11 |
| Apr 16 | 0 | 66 | 465 | 432 | 412 | 11 |
| Apr 17 | 1 | 66 | 465 | 432 | 412 | 4 |
| Apr 18 | 0 | 89 | 421 | 377 | 350 | 0 |
| Apr 19 | 0 | 89 | 377 | 332 | 306 | 13 |
| Apr 20 | 0 | 22 | 399 | 388 | 381 | 0 |
| Apr 21 | 0 | 22 | 377 | 366 | 359 | 0 |
| Apr 22 | 0 | 22 | 399 | 388 | 381 | 0 |
| Apr 23 | 0 | 22 | 310 | 299 | 293 | 0 |

**Table 4:** Predictor set balance and RMSPE for figure 1

|  | e(X_balance) |  |
| --- | --- | --- |
|  | Treated | Synthetic |
| inzidenz_7(2) | 155.14 | 154.25 |
| inzidenz_7(7) | 88.65 | 89.88 |
| inzidenz_7(12) | 66.49 | 68.39 |
| inzidenz_7(17) | 243.79 | 241.99 |
| inzidenz_7(22) | 332.45 | 332.44 |
| inzidenz_7(27) | 221.63 | 222.83 |
| inzidenz_7(31) | 332.45 | 329.46 |
| depr_young | 21.00 | 20.72 |
| depr_old | 53.70 | 49.88 |
| female | 50.40 | 50.29 |
| popdens | 194.00 | 176.93 |
| type | 50.00 | 46.68 |
| incomm | 72.50 | 68.77 |
| outcomm | 85.60 | 80.68 |
| accessibility | 21.00 | 30.99 |
| pharmacy | 39.00 | 49.19 |
| transport | 96.00 | 87.20 |
| ratioemp | 44.60 | 47.45 |
| RMSPE | 28.90 |  |

**Figure 1:**
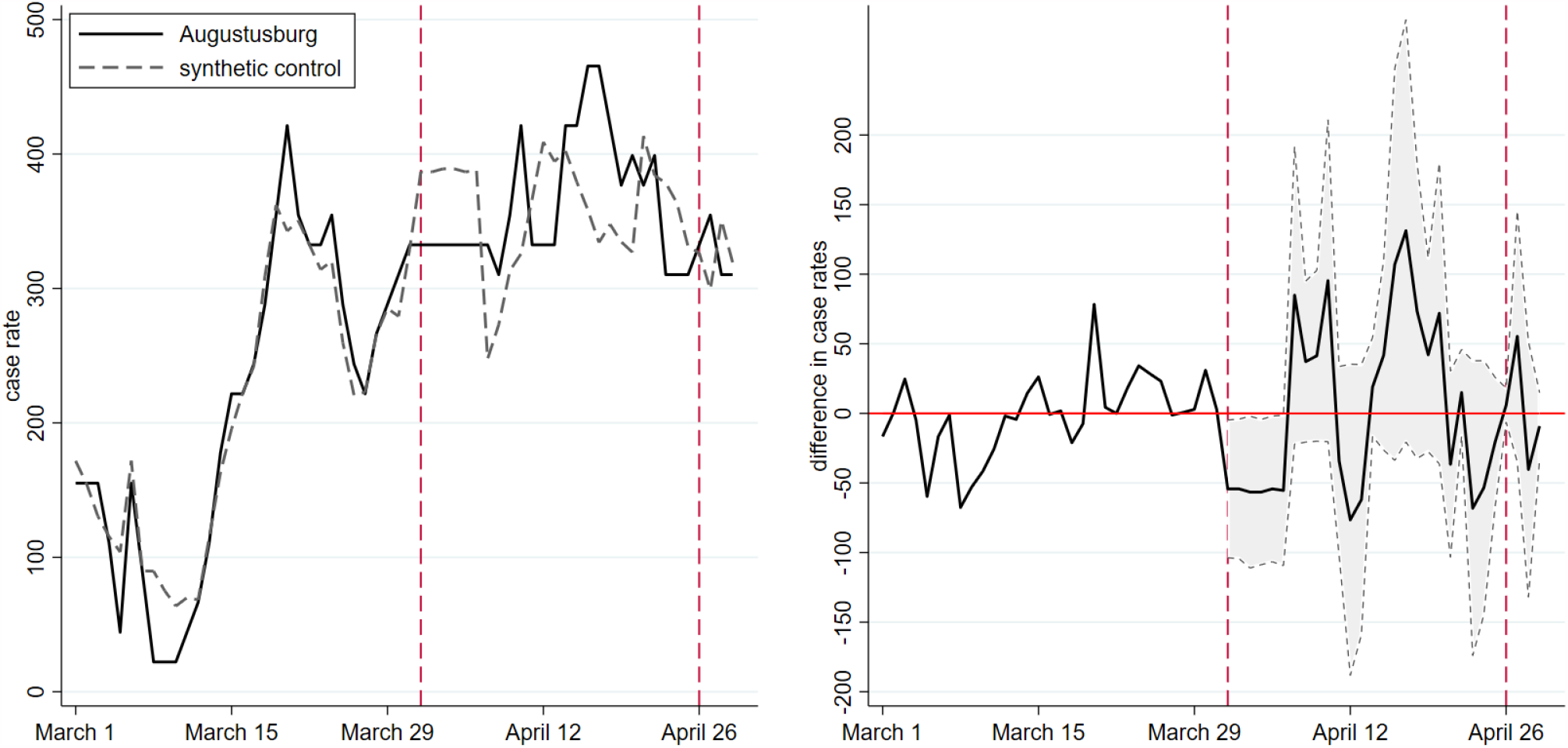
Augustusburg and its synthetic twin compared by case rates

The left panel of the figure shows the pre-treatment fit between Augustusburg (solid graph) and its synthetic twin community (dashed graph) before April 1 (red vertical line) and the effect of T&O thereafter. The pre-treatment fit is very smooth despite the higher volatility of case rates in communities (compared to counties) given their smaller population size. The same increase in the number of infections has a larger effect on rates in smaller communities than in larger counties. The good fit between Augustusburg and its synthetic twin before April 1 (their graphs almost lie one above the other) means that the pandemic in the control community evolved in a very similar way as in Augustusburg.

The evolution of Augustusburg and its synthetic control is less synchronized after April 1 - as one would expect. What is surprising is that differences of case rates are only temporary. On average, case rates evolved in the same way in Augustusburg as in its control community. T&O had no negative public health effect.

The right panel makes this point even more strongly. It shows the difference between Augustusburg and its twin plus confidence intervals at the 90% level. We see that the difference basically fluctuates around zero. The lower bound of the confidence interval, shown by the grey area, is never above the horizontal axis. By this standard, T&O did not lead to significantly higher case rates. Note that we obtain this finding *without* taking the effect of more reported cases due to more testing into account. We will turn to this effect in the discussion section.

## 2 Method

We estimate the causal effect of T&O (the ‘treatment’) on infection dynamics in Augustusburg (the ‘treated unit’) by comparing it to a synthetic control unit. This ‘synthetic twin’ needs to be comparable in terms of relevant socioeconomic factors as well as in terms of pre-treatment trends. We employ the synthetic control method (SCM), proposed for the causal assessment of policy interventions on the basis of aggregate outcome measures, see [1, 2, 3, 4]. This method is the vehicle for our empirical identification strategy.

The method’s core consists of an estimator which identifies communities in Saxony which are comparable to Augustusburg. This comparison is based on information observable prior to treatment and summarized by a set of predictor variables. Table 4 displays the full list of predictors for our baseline scenario.

SCM requires an a-priori list of communities from which to construct the control unit (the ‘donor pool’). Our donor pool consists of all communities in Saxony, except for Augustusburg and communities with a distance of less than 5 kilometers as shown in figure 5.

**Figure 2:**
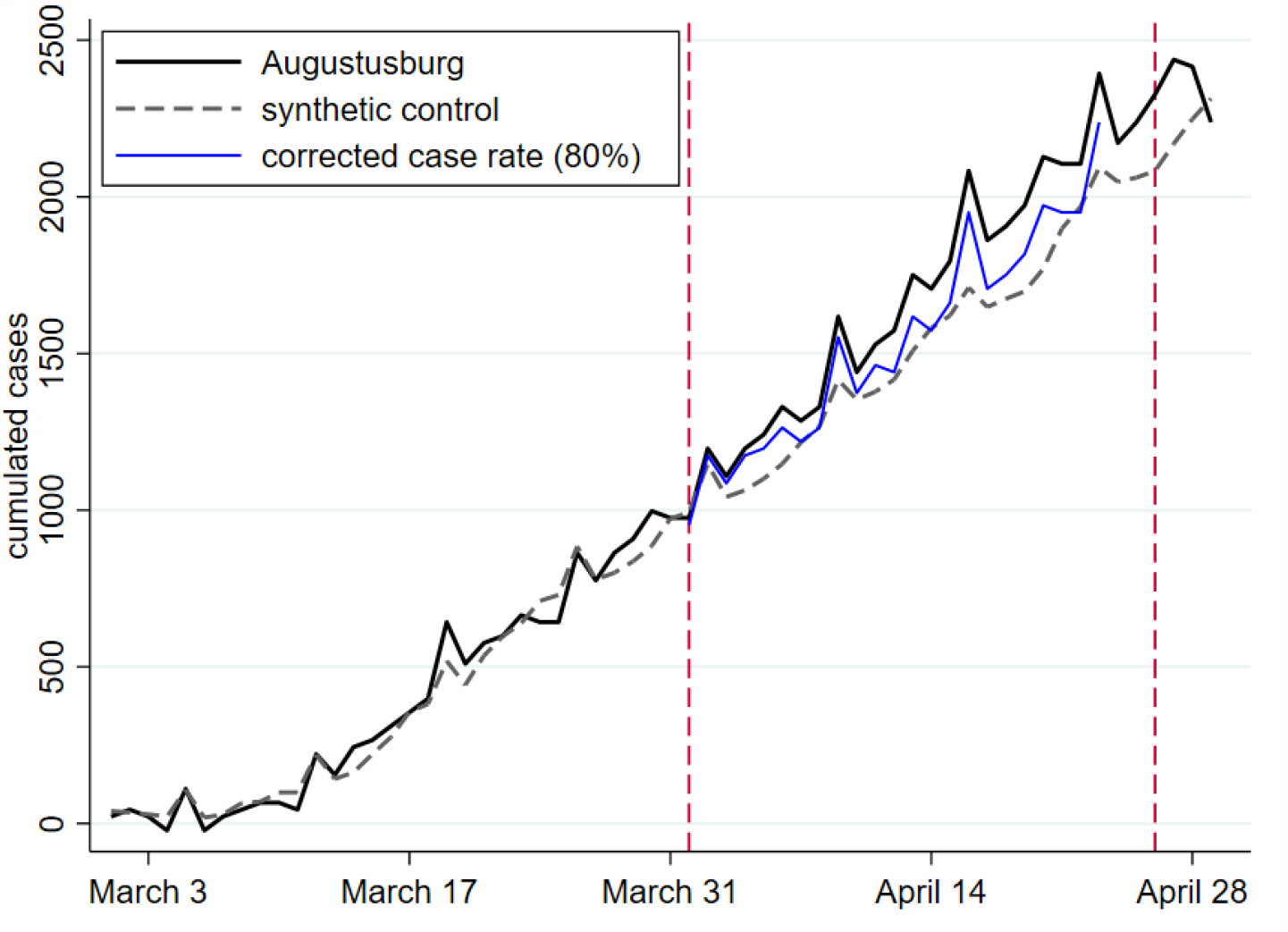
Augustusburg and its synthetic twin compared by cumulative cases

**Figure 3:**
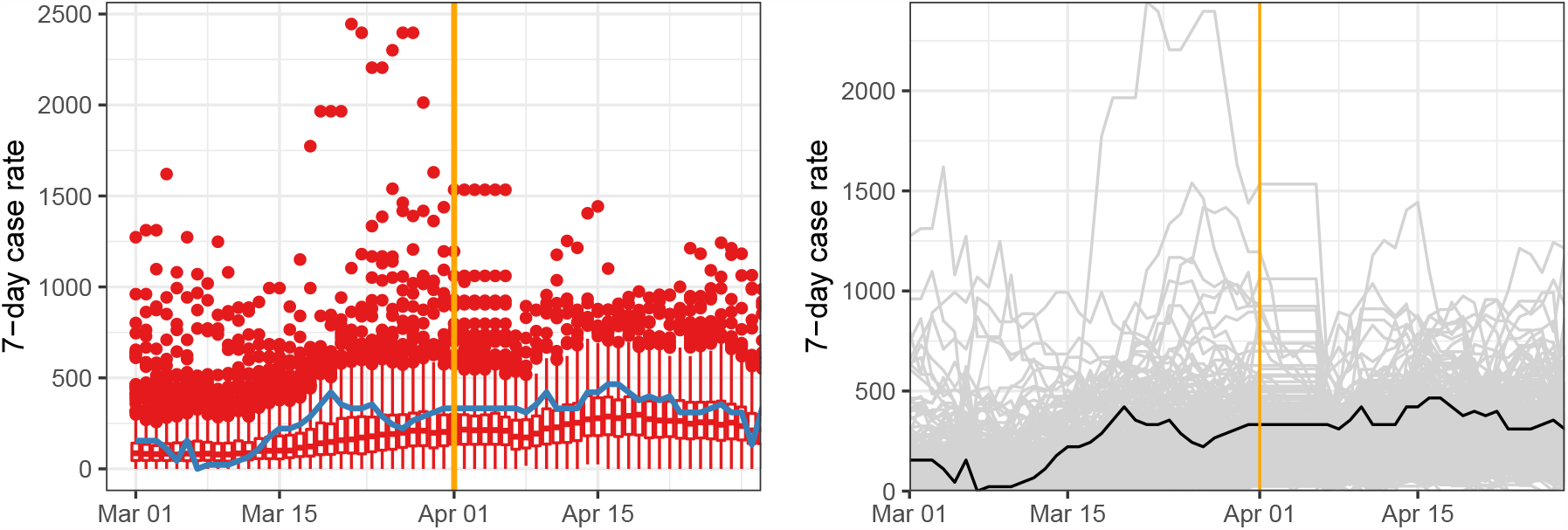
Case rates in Augustusburg (blue) and all other Saxon municipalities (red): Box Plots left and line chart right

**Figure 4:**
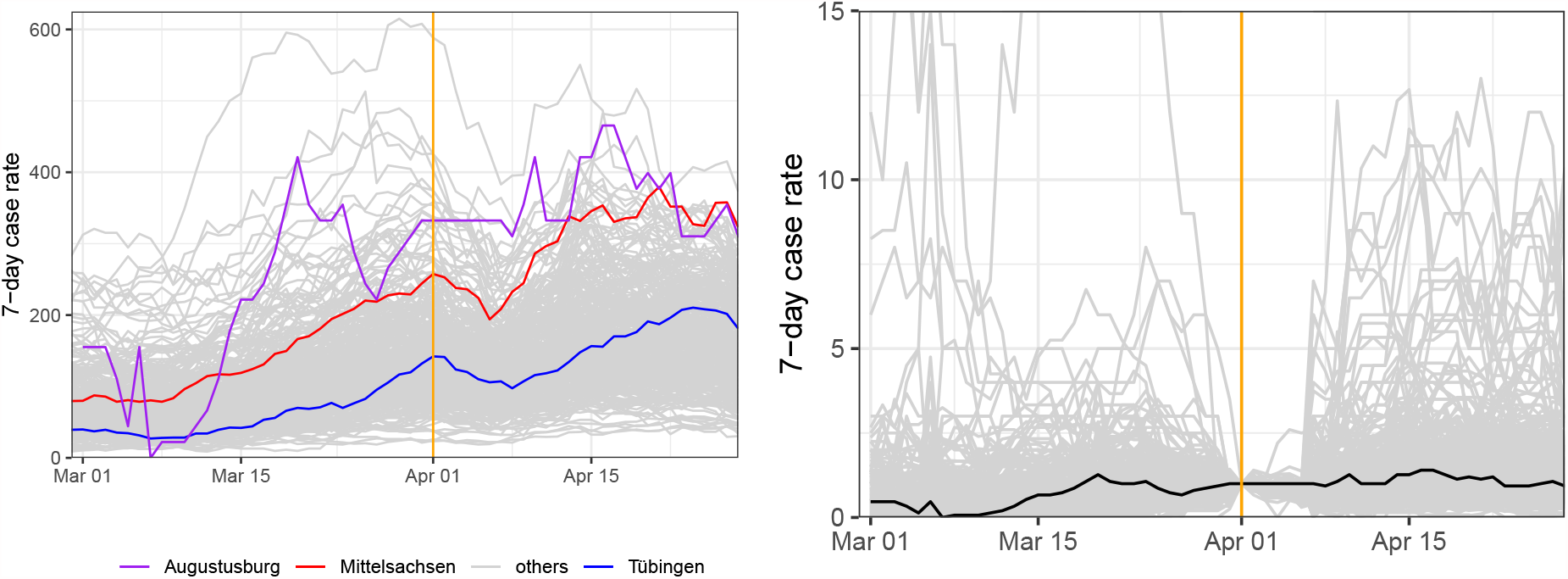
Case rates in Augustusburg, two selected counties and all other German counties (left) and case rates in all Saxon municipalities normalized on the treatment date: Line charts

**Figure 5:**
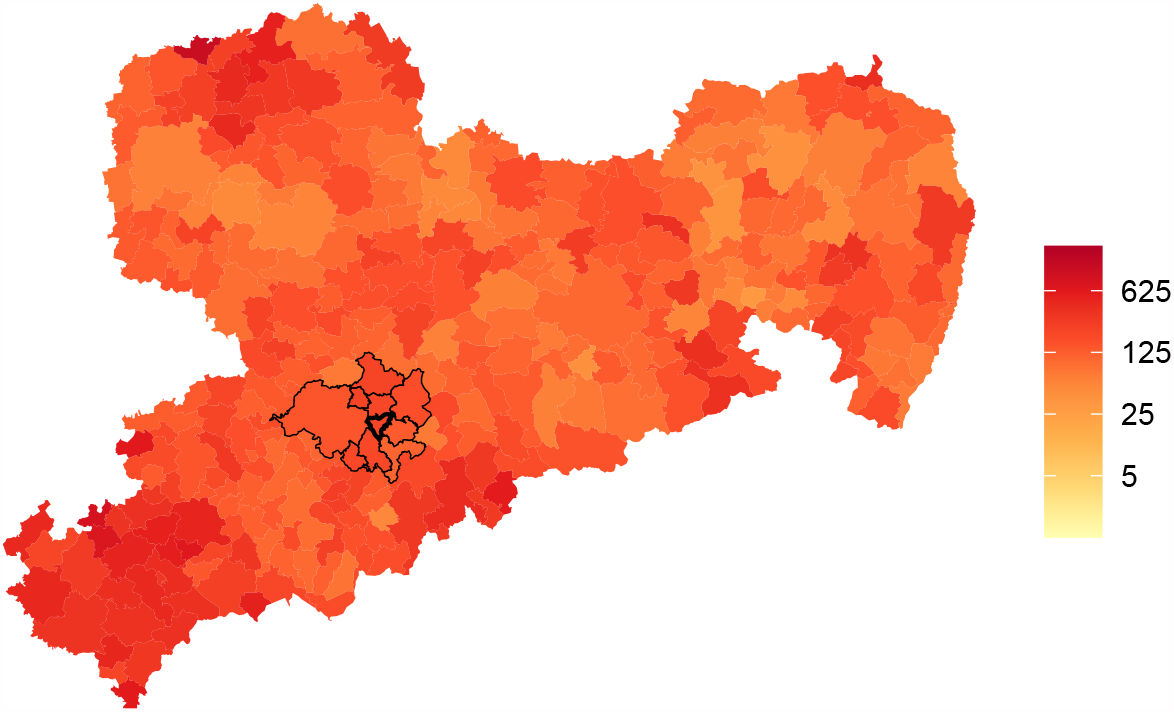
Average pre-treatment case rates in all Saxon municipalities

SCM has been used successfully for evaluation before. Examples include the analysis of Brexit in [5] and the beneficial effects of face masks in [7]. The application of SCM which comes closest to our study of Augustusburg is the analysis of the T&O experiment that started in Tübingen on March 16 in [6]. The SCM approach has also been used in the interim evaluation of the Liverpool mass-scale testing project [9]. This pilot was centered around repeated testing of asymptomatic individuals without any benefits for negatively tested individuals.

## 3 Discussion

Basing a conclusion on one approach only would put a lot of trust in the assumptions underlying this one approach. We therefore undertook various robustness checks. First, we changed our pandemic measure. Instead of looking at case rates, we measure the pandemic by the number of cumulative cases since March 1. Second, we changed the donor pool for our baseline analysis by removing one community of our synthetic control unit shown in table 1 one by one.

### 3.1 The effects of testing and opening based on daily cases

We now study the effect of T&O by adding up the number of cases that occurred in Augustusburg and in all other communities in Saxony since March 1. This is a more ‘severe’ test of infection effects: in seven-day case rates, additional infections are no longer counted after 7 days. When adding up all cases since some starting point, an extra case is always present in the pandemic measure.

- The plain effect Figure 2 shows that the pre-treatment fit before April 1 is again very good, even better than in our baseline figure. This is not surprising as adding up numbers since some fixed starting point is a less volatile process than adding up number over a period of the previous 7 days only. As before, Augustusburg and its control community basically follow identical pandemic paths before April 1. The control regions resulting from fitting pre-treatment infection dynamics in addition to other criteria (see again the predictor balance in table 6) and their weights are shown in table 2.

**Table 5:**
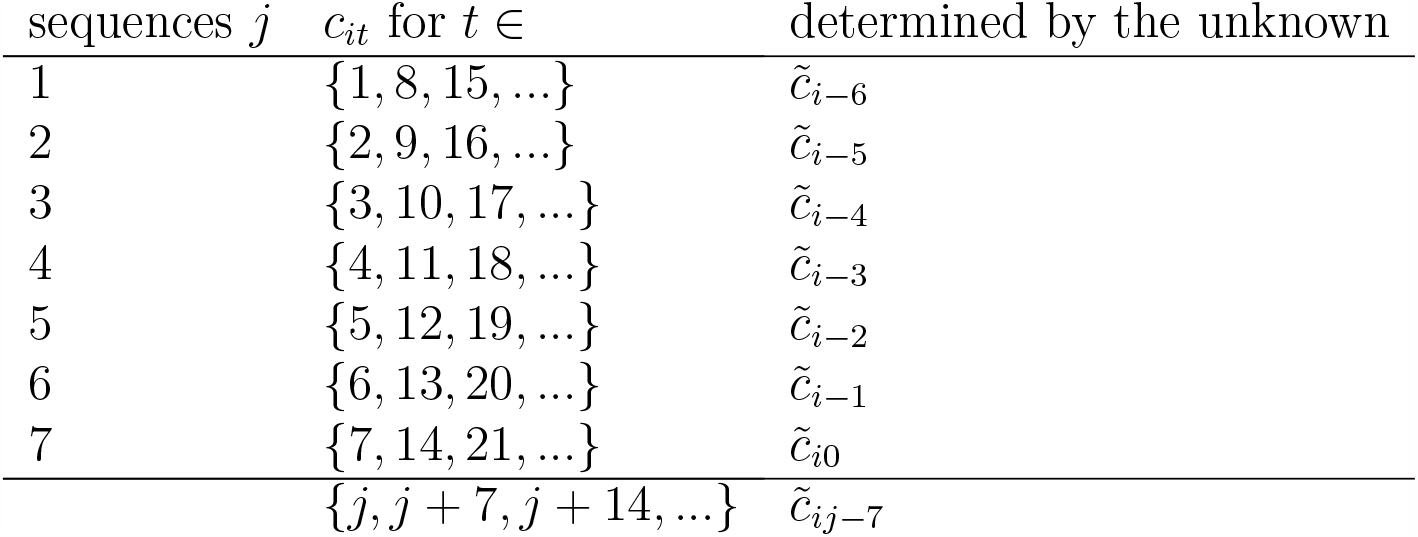
Auxiliary variables 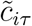 and their effects

**Table 6:**
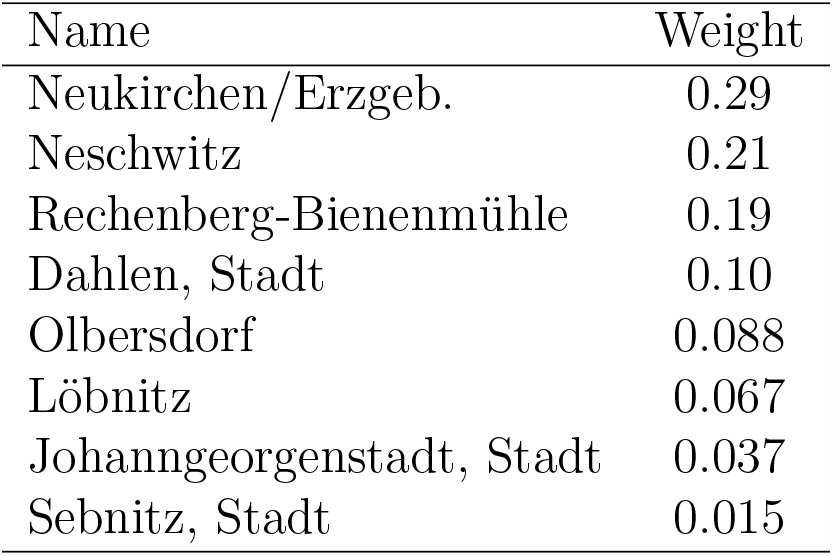
Predictor set balance and RMSPE for figure 13

Turning to the treatment effect of T&O for the time after April 1, Figure 2 shows that the difference in the total number of infections between Augustusburg (solid graph) and its synthetic twin (dashed graph) moderately increases over time. It then comes at no surprise that the absolute Augustus-twin difference, shown in the right panel of figure 13, never significantly differs from zero. The percentage difference, shown in figure 12, ranges between 0% and 20%. The overall effect at the end of the T&O period is 10% only.

**Figure 6:**
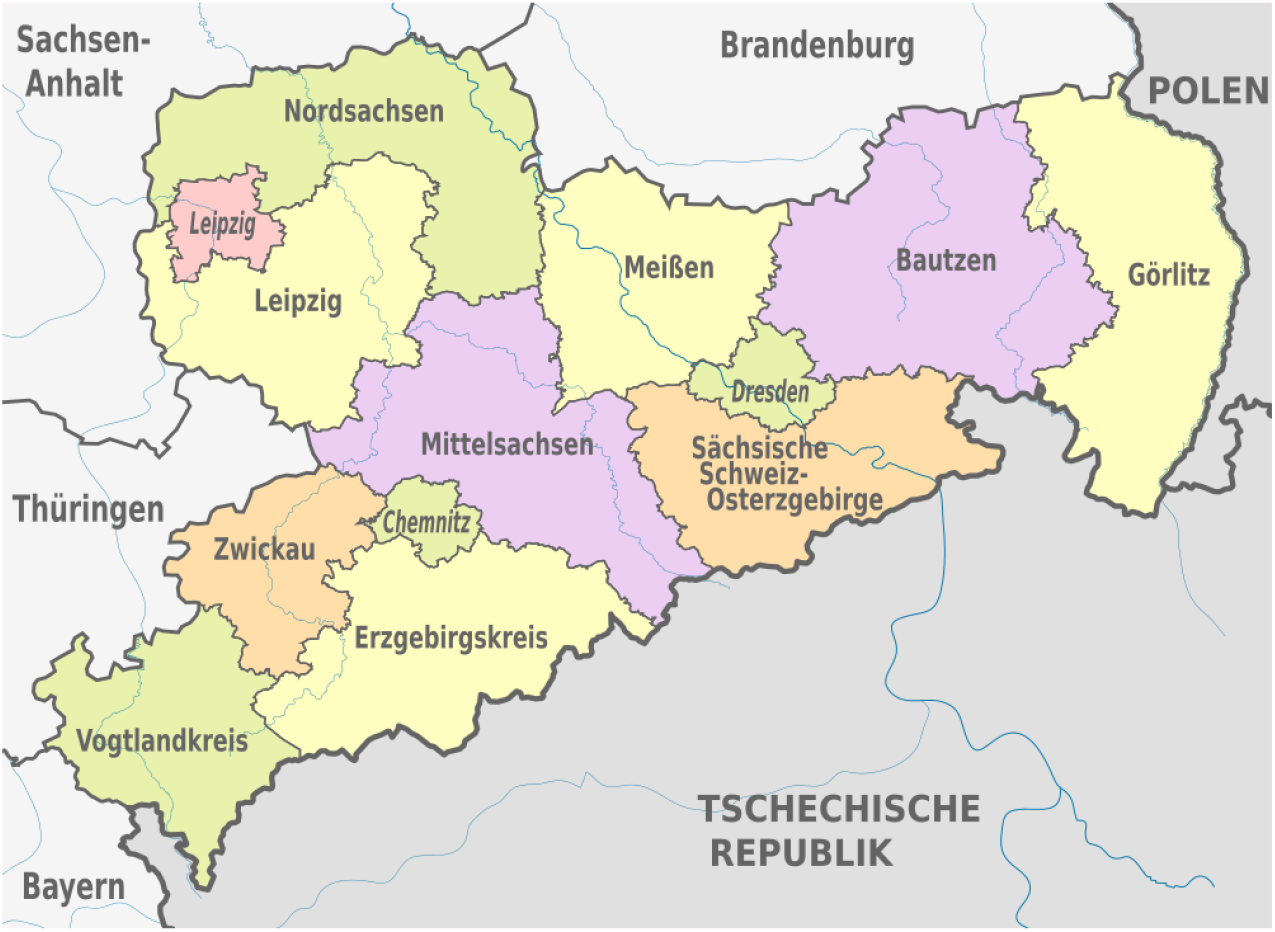
The counties (10 ‘Landkreise’ and 3 ‘kreisfreie Städte’) in Saxony - by courtesy of Wikipedia

**Figure 7:**
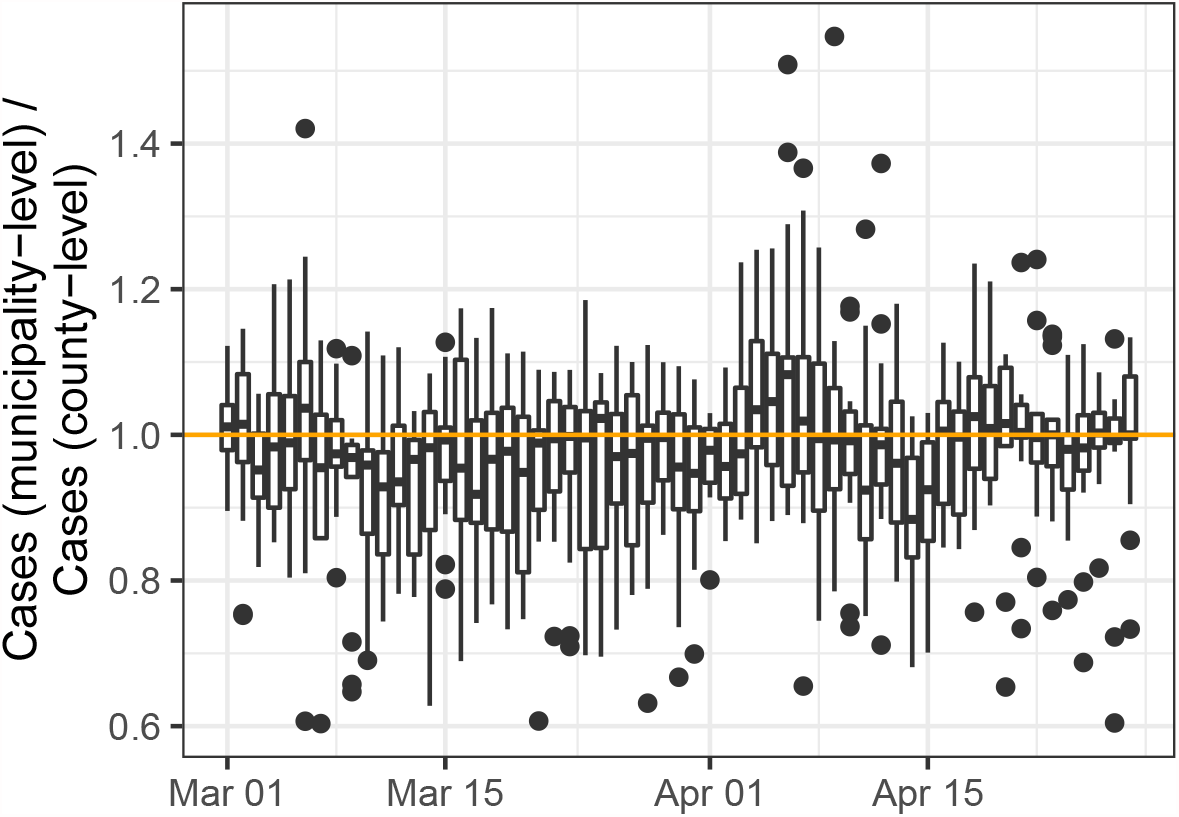
Comparing case rates published by RKI and by communities in Saxony

**Figure 8:**
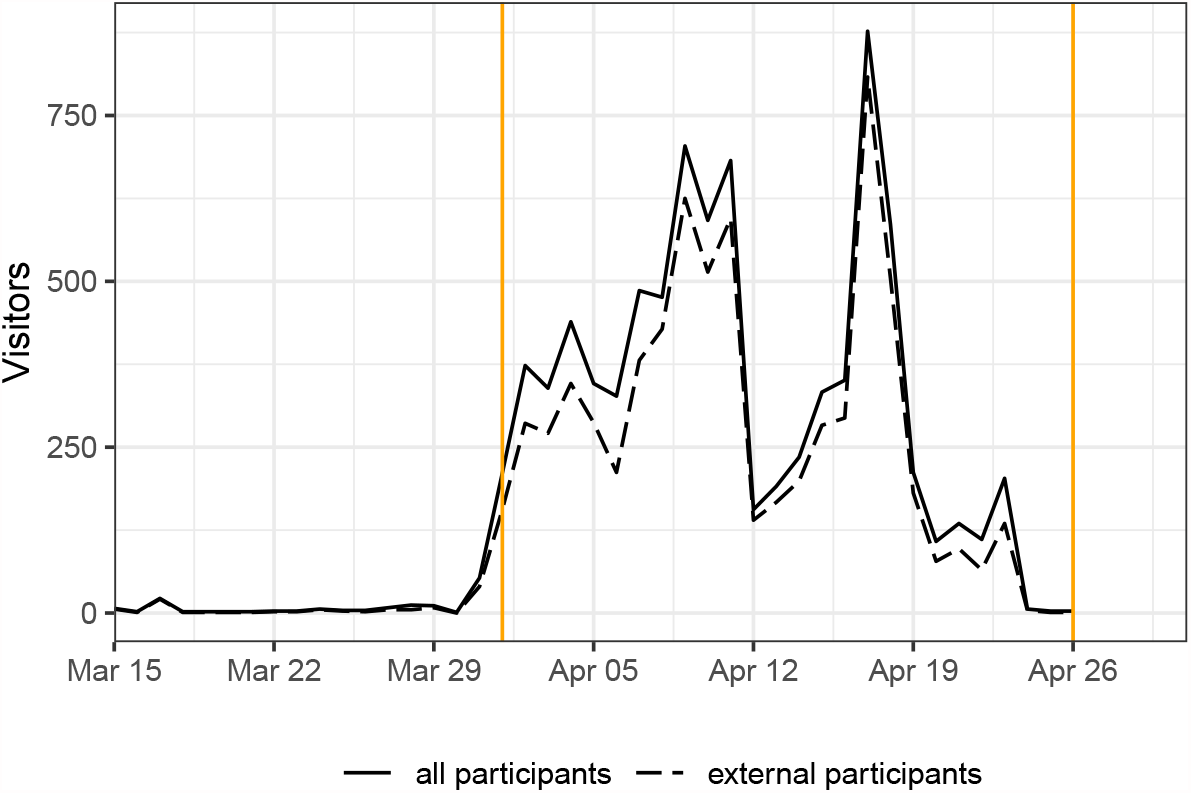
Daily visitors to Augustusburg

**Figure 9:**
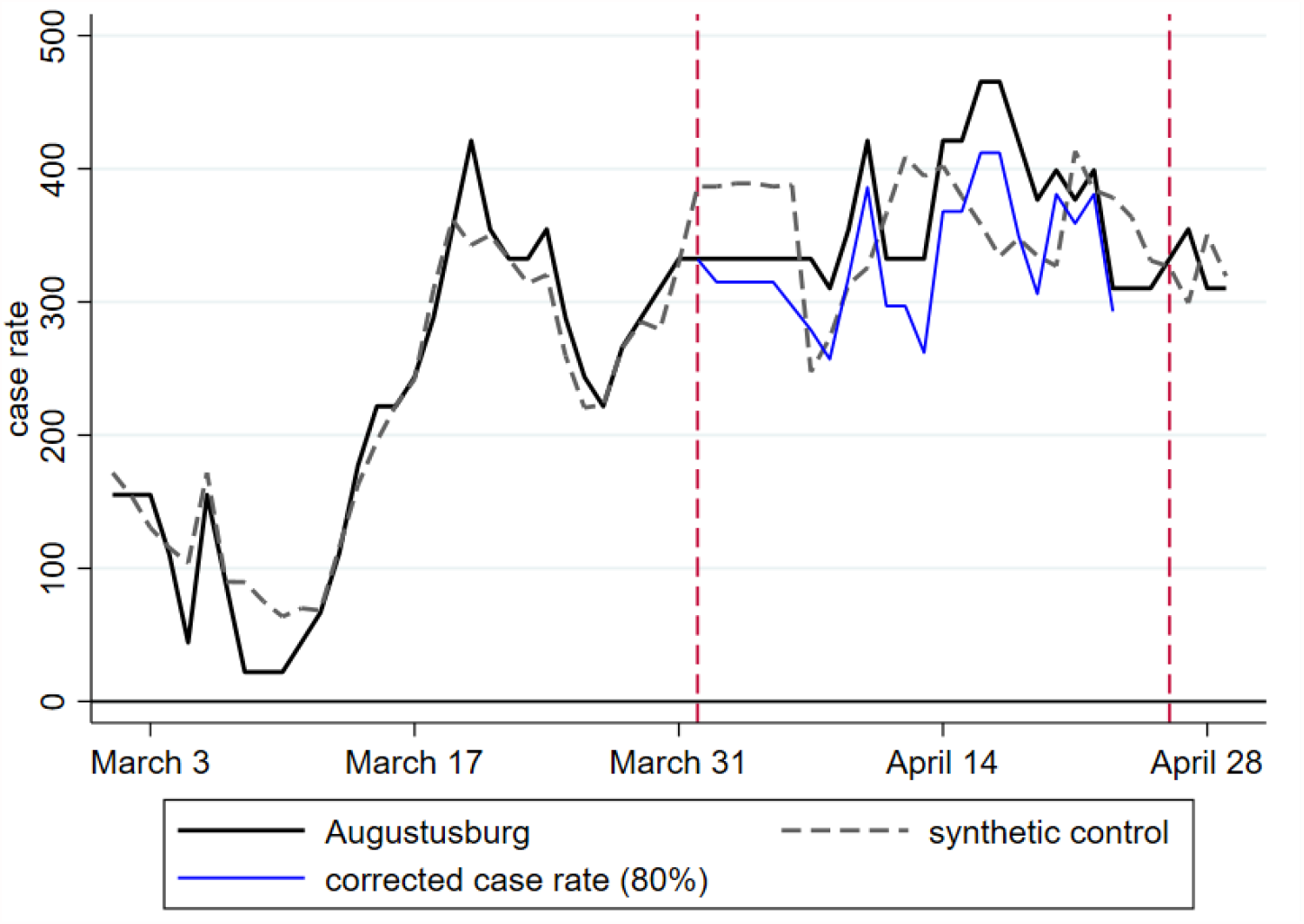
Augustusburg, its synthetic twin community and corrections for rapid testing

**Figure 10:**
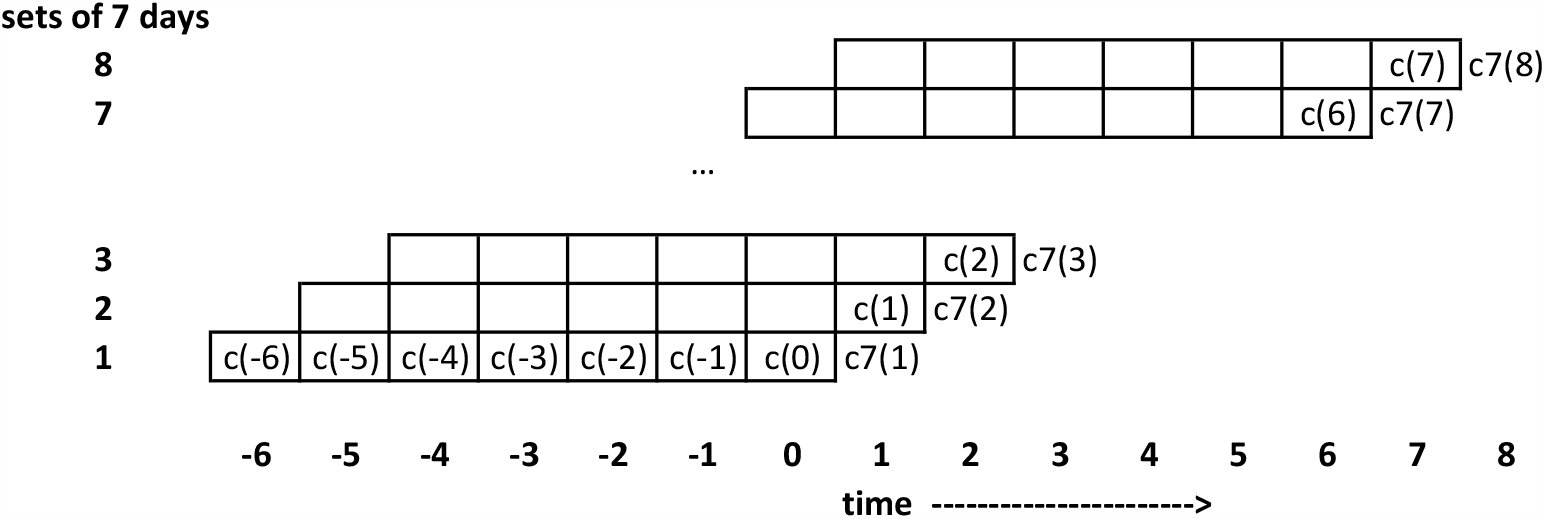
Recovering daily cases from case rates *Note:* Daily cases c(−6) to c(0) are unobserved and are denoted 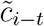 in the text.

**Figure 11:**
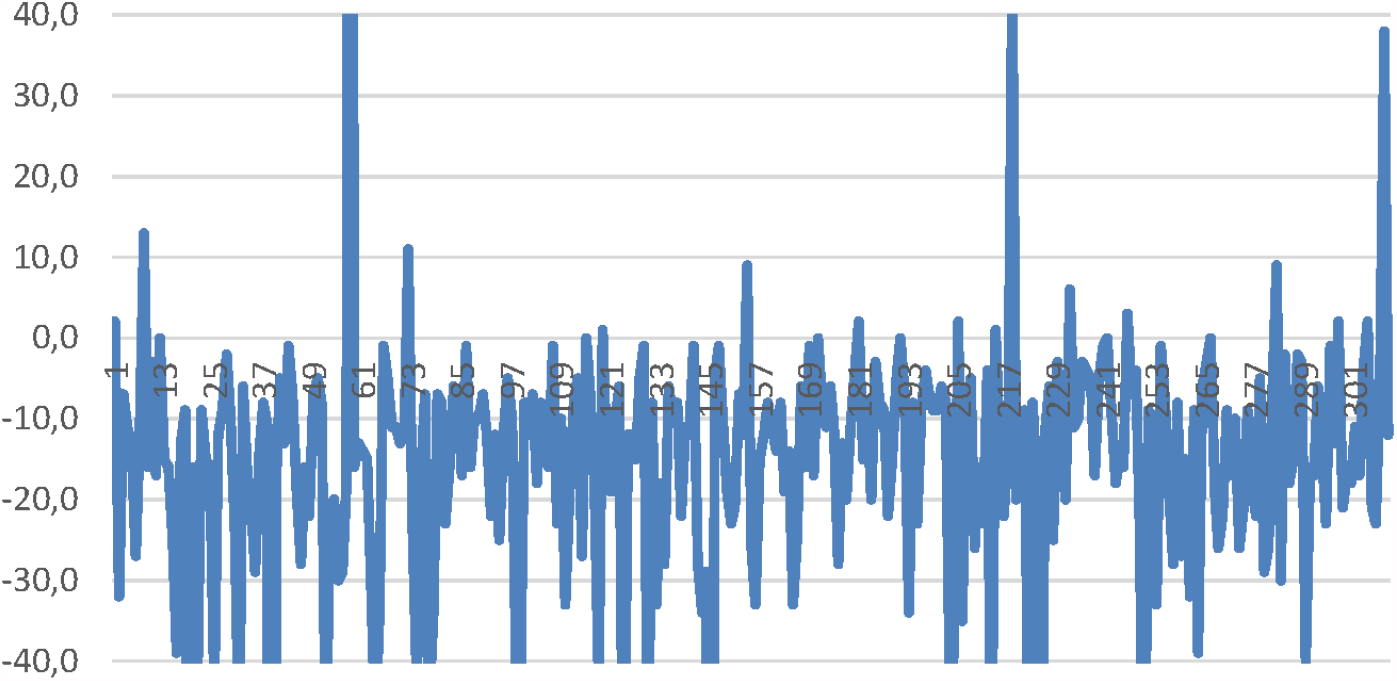
The sum of minima over weekdays by community

**Figure 12:**
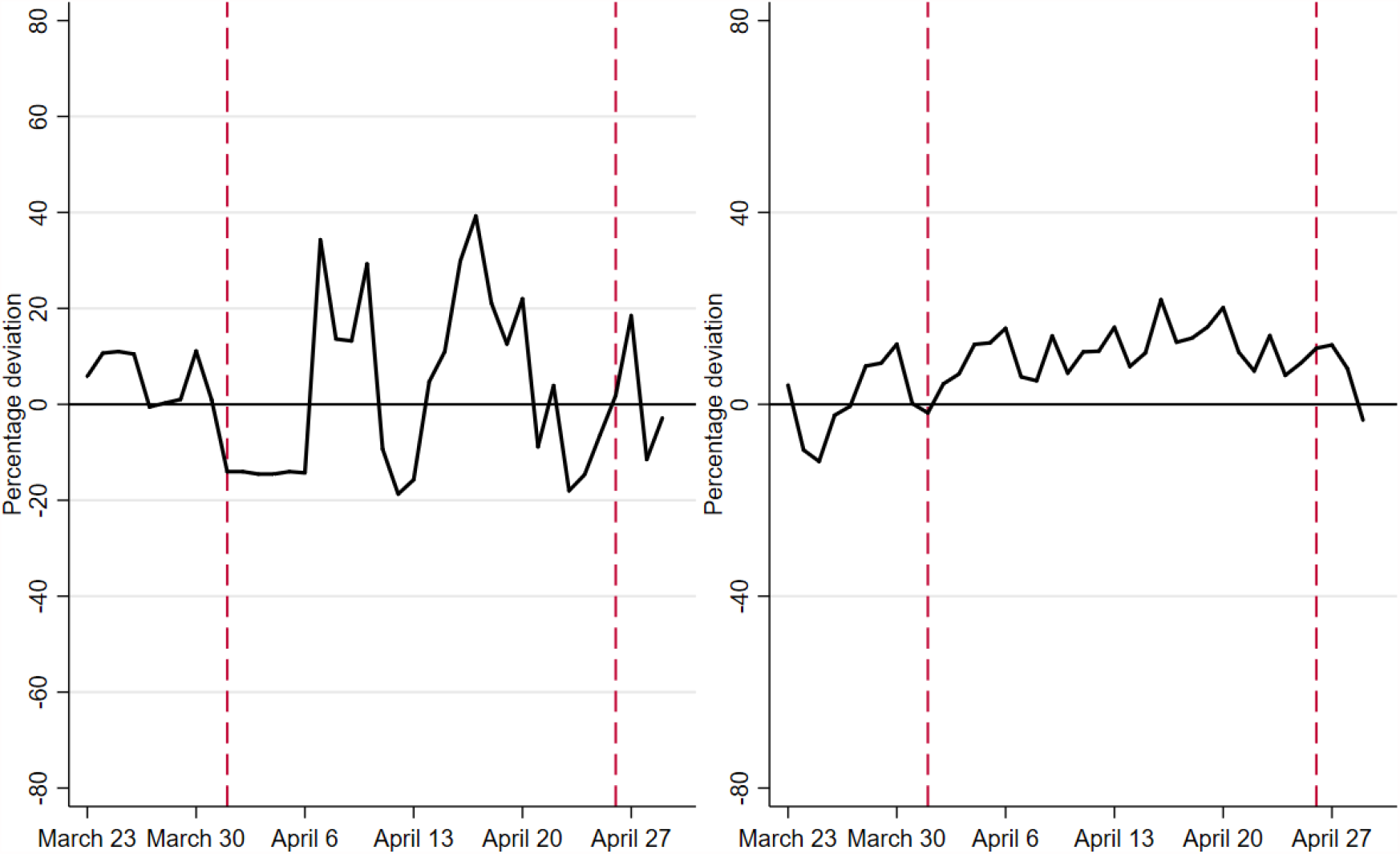
Percentage deviations of case rates (left) and cumulative cases (right)

**Figure 13:**
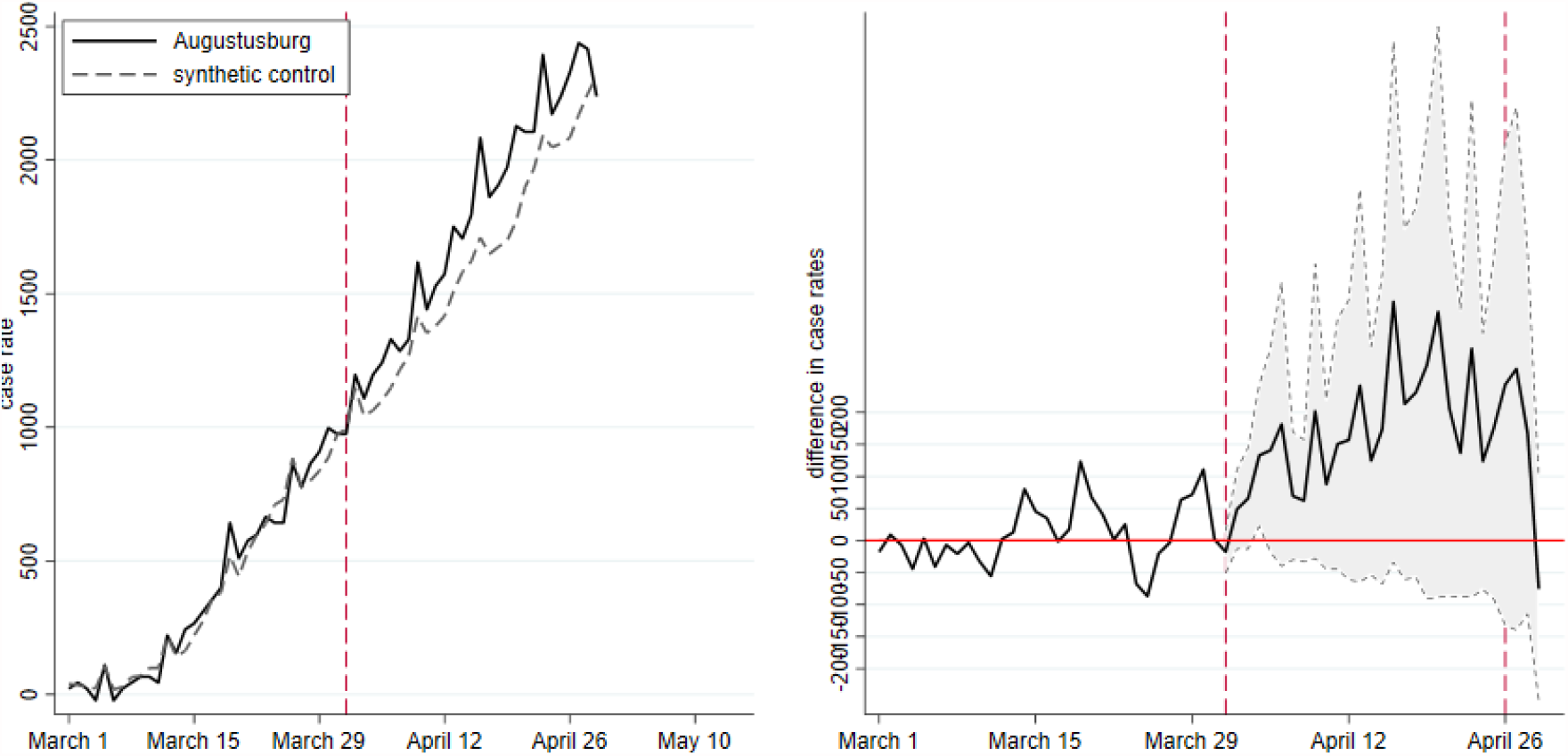
Augustusburg, synthetic twin, cumulative cases and confidence interval

- Data issues (again) We should not ignore, however, that the graphs in figure 2 are theoretically impossible. When we add up the daily number of cases since March 1, the graph should never fall: a negative number of daily cases does not exist. Yet, our method, based on officially reported case rates do imply negative numbers. As we believe that this is a measurement problem that is identical across all communities in Saxony and is not specific to Augustusburg, we still believe that our results are informative.
- Correcting for the effect of testing Our findings so far have shown that T&O as implemented in Augustusburg did not have a strong, if any, effect on infections. We can now go one step further. T&O implies more tests and more contacts. More tests help to identify infectious individuals. Opening implies more contacts. The former implies a dampening effect for the pandemic, the latter makes the pandemic stronger.

More testing not only identifies infectious individuals, it also leads to more reported cases in the data (provided that a positive rapid test is confirmed by a PCR test). In order to take this effect on official cases or case rates into account, we start from the number of positive daily tests, reported in the first column of table 3. In a world with perfect data, one would know how many of these tests are PCR confirmed. Only PCR confirmed tests are reported to the health authorities and enter official statistics. Table 3 assumes that between 50% and 80% of positive tests would be PCR confirmed.

When we subtract 80% of the number of positive tests from the cases observed in Augustusburg, we get an estimate of the cases that would have occurred in the absence of a high number of rapid tests. When we plot the corrected cases as the blue graph into figure 2, we see that Augustusburg performs almost as well as its synthetic control. As the full difference without correcting is already insignificant, it is clear that the remaining difference is not statistically significant either.

### 3.2 ’Leave one out’

A standard approach to testing how sensitive results are to changes in the synthetic control community consists in taking individual communities out of the control community. We therefore re-ran the SCM for donor pools that consisted of all communities in Saxony (apart from neighboring communities and Augustusburg itself) minus one of the selected comparison communities visible in table 1.

As figure 14 shows, some communities are crucial. When we leave out Regis-Breitingen Stadt, the community with the largest weight, Augustusburg performs worse relative to its new synthetic twin. When some other communities are left out, Augustusburg performs better. For most communities, obviously for those with low weights, the findings remains within the same range as in the main part. We conclude that ‘leaving one out’ confirms our baseline finding.

**Figure 14:**
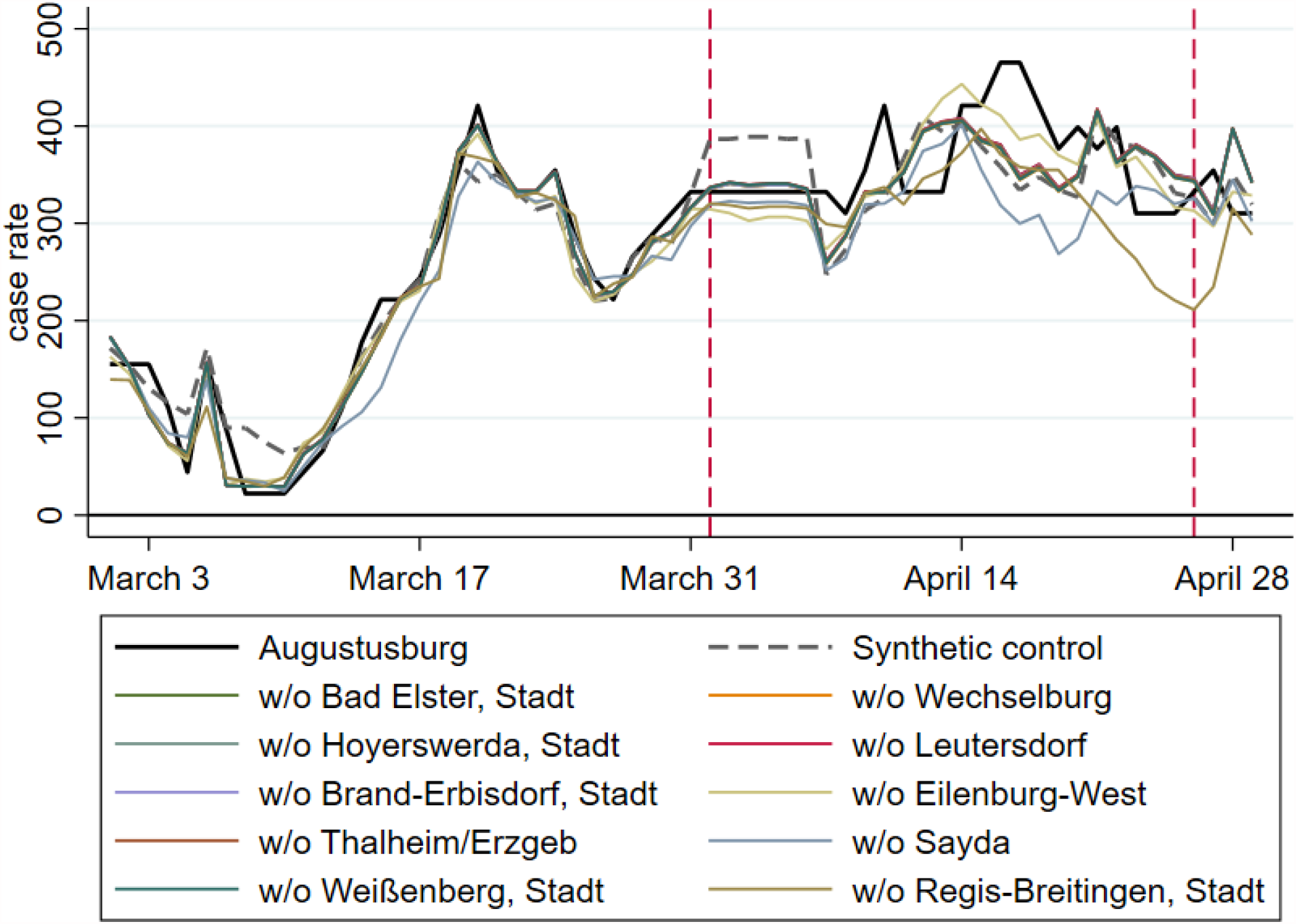
How much does the synthetic control group depend on each of its counties?

### 3.3 Why testing and opening worked so well in Augustusburg

When we compare Augustusburg to Tübingen (see [6] for a detailed analysis of the latter), figure 4 (left) shows that Tuübingen started its T&O with extremely low case rates (compared to other counties) while case rates of Augustusburg were very high. One could therefore argue that Tuübingen had a much higher risk of a bad outcome of T&O as most visitors came from regions with higher case rates.

There are two aspects this argument ignores, however. First, all visitors, both in Tuübingen and in Augustusburg, require a negative test. Whether visitors come from high incidence or low incidence regions, they are in principle identical – all negatively tested. Second, our results that Augustusburg performed so well is based on the performance of Augustusburg relative to other communities in Saxony. Compared to these peers, Augustusburg was in the middle range, as shown by figure 3 (right). Augustusburg could therefore also have experienced an increase in case rates.

What about the number of visitors? If Tuübingen experienced more visitors relative to population size than Augustusburg, one could also conjecture that the increase in incidence in Tuübingen is due to the higher number of visitors. The number of visitors to Augustusburg from outside lay at around 6% of its population size (figure 8) per day. In Tübingen, the number of visitors from outside Tuübingen cannot be identified from available data. The total number of participants as measured by the number of tests relative to population size of Tuübingen city is around 4%. The number of visitors from outside Tuübingen is therefore lower than 4% and, in any case, lower than the (relative) number of visitors to Augustusburg. Hence, visitor (over-) flows cannot explain the relative difference between Augustusburg and Tübingen either.

What speaks in favour of Augustusburg is its very stringent surveillance system. Participants in T&O were not only tested initially, they were tested every day. This might have created incentives for visitors to behave carefully. Augustusburg also registered all visitors in every single activity that belonged to T&O. When entering a restaurant or a museum, visitors were registered. They were de-registered when they left. This might have strengthened the feeling for the importance to respect hygiene rules. It also made sure that no untested persons could participate in events.

Despite data quality issues and a potential ‘advantage’ from having already had a high case rate at the beginning of T&O, we conclude from our preliminary findings that T&O worked very well in Augustusburg. We cannot identify a significant increase due to T&O, neither in case rates, nor in cumulative numbers of cases. We hope that we can resolve data problems in the weeks to come and analyse individual data. At this point, we see no reason why T&O had to be stopped end of April.

## Data Availability

public data

## Acknowledgments

We would like to thank a long list of colleagues for inspiring discussion, comments, criticism and encouragement. We are especially grateful to René Glawion, Gernot Muüller, Peter Kremsner and Dominik Papies. Authors declare no competing interests. No financial support has been received by any of the involved or evaluated parties.

## A Appendix

### A.1 Data

#### A.1.1 General Information

We acquire data on cases and case rates on two different levels of geographic detail. Because it allows for a comparison between communities, we rely on case rate data from the Saxon State Ministry for Social Affairs and Cohesion [15] in our central analysis. We observe case rates beginning at the start of the experiment on April 1 2021 up to its end on April 23. To enable comparisons, we incorporate data starting on March 1, 2021, one month before treatment.

To achieve the control group most suited for our analysis, we introduce a range of time-invariant predictors on demographic structure and regional differences in health case systems, which we acquire from Inkar [12], in its most recent available version from 2017. The following predictor variables are included: old- and young-age dependency rations (in %), the average age of the female population (in years), the population density (in %), the regional settlement structure (a dummy), the share of incoming and outgoing commuters in the local workforce, accessibility indicators, pharmacies per inhabitant, transport indicators and the unemployment rate (in %). As a result from limitations in the data, we combine the smallest of municipalities with their peers according to the official Saxon ‘Verwaltungsgemeinschaft’ and ‘Verwaltungsverband’ grouping [16].

#### A.1.2 Descriptive statistics for communities

At the end of the day, the synthetic control method identifies a certain set of communities in Saxony and compares pandemic dynamics in Augustusburg with these identified communities. To obtain a first feeling about the possible outcomes of this comparison, we first look at the evolution of the pandemic in Augustusburg, measured by the seven-day case rate, and in all other communities. The seven-day case rate, case rate in short, is the sum of new infections over the previous seven days per 100,000 inhabitants, see (A.1).

Figure 3 displays the seven-day case rate in Augustusburg and in all other communities in Saxony. The start date of the T&O experiment on April 1 is highlighted in orange. The distribution of infections in the latter are represented by box plots in the left panel and as individual lines in the right panel. It becomes clear that the case rate of Augustusburg was not exceptional compared to its peers. We should keep in mind, however, that beginning of March, Augustusburg had the lowest case rate in the sample.

While Augustusburg is representative of communities in Saxony, it is at the upper end of the distribution of case rates of German *counties*. A comparison of a community with a county is a bit unfair though, as a cross-section of counties, being larger units, tends to have a lower variance. Communities therefore find themselves at extremes of a distribution more easily than counties. Yet, the left panel of figure 4 shows the relatively high case rates of Augustusburg, also compared to its own county of Mittelsachsen. It is also interesting to see that case rates are considerably higher than in Tübingen county, the second famous opening & testing region in Germany. The right panel of figure 4 shows case rates in Augustusburg compared to all German counties and normalized on the treatment date. It confirms the insight from figure 3 that Augustusburg was representative of Saxony before and after the treatment date. This suggests, *before* employing SCM, that SCM should be able to capture the dynamics in Augustusburg well.

To put Augustusburg in the geographical context of Saxony, Figure 5 plots the average pre-treatment case rates (March 1 to 31) in a map of Saxon communities. Augustusburg and its neighbors, defined as distancing less than 5 km and excluded from the donor pool of SCM analysis, are highlighted with bold borders. These communities are Amtsberg, Augustusburg, Chemnitz, Flöha, Frankenberg, Leubsdorf, Niederwiesa, Oederan, Wildenstein and Zschopau.

One difference of our study compared to earlier CoV-2 analyses in Germany based on SCM (see e.g. [13] or [7]) is our focus on communities (’Gemeinden’) rather than counties (’Landkreise’). It is therefore of obvious importance to understand how well the data at the community level is consistent with data at the level of the county. We therefore add up cases in all communities of the county and compute the ratio relative to county cases. The result is shown in figure 7.

#### A.1.3 Descriptive statistics for individual data

Figure 8 shows the number of participants in T&O per day. We equate the number of external participants with the number of tests by individuals with a postcode other than Augustusburg.

Only 17% of all participants throughout the timeframe were Augustusburg locals. As the figure shows, the number of visitors to Augustusburg amounted to around 306 per day between April 1 and 23. It ranged from 65 on weekdays to 808 on weekends. Given its 4,512 inhabitants, the average number of visitors to the city amounted to 6% its population per day. Also including locals, the share of participants lies at 8% of its population.

The second column of table 3 displays the number of positive rapid tests by inhabitants of Augustusburg per day. The numbers were collected and reported by Theed. The corresponding seven-day case rate is in column three. It is computed as seven-day case rates for infections as in (A.1).

### A.2 Literature

The effect of testing and opening has been analysed in parallel e.g. for the city of Tübingen. The literature section here is therefore relatively brief and we refer readers to [7].

The desirability of mass testing has been emphasized by many commentators. A theoretical analysis showing that testing and quarantining can dramatically reduce the costs of an epidemic is in [8]. The empirical merit of T&O is largely unstudied [11].

SCM has been frequently used in the social sciences to study the effect of policy interventions, broadly defined, on political, social, and economic outcomes [1]. In these contexts, SCM has been shown to be a flexible and robust estimation tool. In addition, it has also been applied to COVID-related research, for instance, to study the effectiveness of lockdown measures by means of a counterfactual analysis for Sweden [4, 6] and to study the effect of shelter-in-place policies in California [9]. In addition, [14] use SCM to study the effect of face masks on SAR-CoV-2 cases in Germany.

### A.3 Findings

Table 4 shows our predictor set employed for the case rate analysis displayed in figure 1.

### A.4 Methods

#### A.4.1 The SCM

We construct a synthetic control unit consisting of the donor pool of comparison regions and by comparing the outcomes of the treated unit and the synthetic control unit after the start of the treatment. The match between treated regions and the synthetic control group is done through a minimum distance approach for a set of predictor variables evaluated along their pre-treatment values for the treated region and those in the donor pool. This ensures that pre-treatment differences in trends of the outcome variable are leveled.

In our application, we make sure that the control unit tracks the outcome variable in Augustusburg during 15 days prior to April 1 as closely as possible. Formally, we construct the control unit by selecting weights on the communities in the donor pool for which we obtain the best match between the control unit and Augustusburg given our control variables. In addition we target socioeconomic characteristics as visible in our lists of all target variables in table 4.

Our implementation follows largely [14] or [7]. We conduct all SCM estimations in Stata using the SYNTH [2] and SYNTH_RUNNER [10] packages.

Confidence intervals (CIs) are calculated from one-sided pseudo *p*-values obtained on the basis of comprehensive placebo-in-space tests. The latter tests calculate pseudo-treatment effects for all regions in the donor pool treating each of the regions as if it would have also re-opened public life and the local economy on April 1, 2021. One-sided pseudo *p*-values are then calculated of the share of placebo-treatment effects that are larger than the observed treatment effects for treated regions and thus indicate the probability that the increase in the number of SARS-CoV-2 infections was observed by chance given the distribution of pseudo-treatment effects in the donor pool.

To account for differences in pre-treatment match quality of the pseudo-treatment effects, only donors with a good fit in the pre-treatment period are considered for inference. Specifically, we do not include placebo effects in the pool for inference if the match quality of the control region, measured in terms of the pre-treatment root mean squared prediction error (RMSPE), is greater than 10 times the match quality of the treated unit [5]. Based on the obtained pseudo *p*-values we calculate confidence intervals as described in [3].

#### A.4.2 Case rates, comparisons and growth rates

Some or our arguments require a little bit of algebra. Especially the derivation of daily cases from case rates in not entirely trivial.

- The basics We start by defining *c*_*it*_ as the number of new cases on day *t* in region *i*. Let *N*_*i*_ denote the population size of region *i*. This allows us to compute the sum of cases over the last seven days as 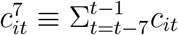 and the seven-day CoV-2 case notification rate per 100,000 (the case rate) as

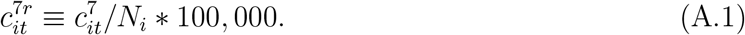

This expression is shown everywhere in this paper whenever we display ‘case rate’ on the axes of the figures or write about seven-day case rates.

- Comparing regions It is sometimes useful to see the absolute difference in cases between Augustusburg and its synthetic control community. The daily difference is

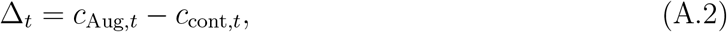

where the number of cases in the synthetic control county is a weighted sum of members *m* of the synthetic control county,

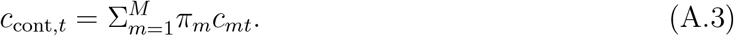

The weight *π*_*m*_ of member *m* of the control county is given by the outcome of SCM.

The difference per week, i.e. over the previous seven days, is

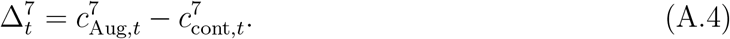

Defining

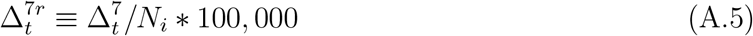

yields the expression shown in all figures with a panel B where the axis is labeled by ‘differences in case rates’.

- Positive rapid testing and case rates Imagine we have data on cases discovered via rapid testing. We denote these cases by 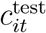 Cases that would have been reported if rapid testing had not taken place can then be approximated by 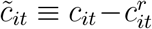 How can we approximate the seven-day case rate, being based on positive PCR tests, that would have been observed in the absence of rapid testing? The seven-day case rate is defined above in (A.1). The number of positive rapid tests over a period of seven days (the weekly number of positive tests, simply speaking) is given by 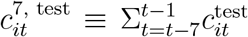.Hence, assuming that each positive rapid test is confirmed by a positive PCR test, the corrected case rate is given by

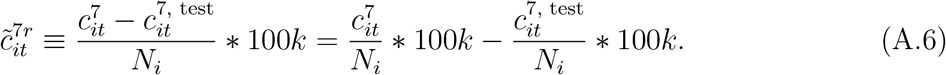

As the equality sign shows, we can either correct cases and then compute the rate or compute the difference between a case rate and a “positive-test” rate.

- From case rates to cases - the principle We also want to use cumulative cases as a measure of the pandemic. To this end, we need cases and not only case rates. We start from case rates 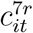 and compute cases *c*_*it*_ as follows.

The case rate is defined in (A.1), 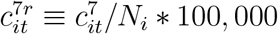. When we solve this for the sum of new cases over the previous 7 days, i.e. for 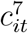, we get

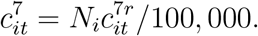

As 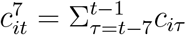 by definition, we can compute

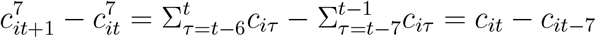

and therefore

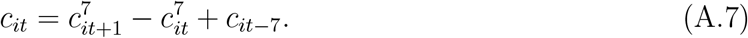

Now let time start on day *t* = 1 as shown in figure 10. We define *t* = 1 as the day where we observed the first seven-day case rate, i.e. 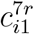. Then, from (A.7),

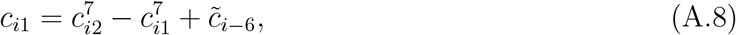

where 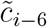 is the (unobserved) number of cases on day −6. For *t* = 2 to *t* = 7, we get

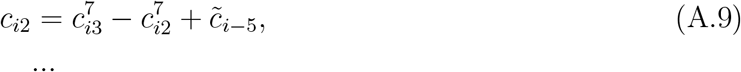

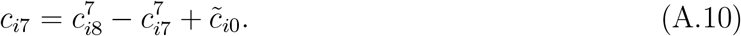

Hence, we could compte daily cases *c*_*i*1_ to *c*_*i*7_ if we knew 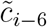 to 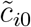. We will turn to the issue of fixing the unknown values in a moment.

Computing daily cases as of *c*_*i*8_ employs (A.7) and yields for *t* = 8

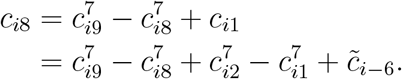

For any 15 *> t >* 7, we get

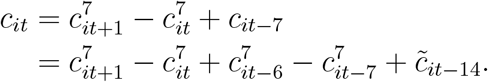

This shows that the assumption about 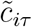 affects all future recovered cases *c*_*it*_.

Keeping this in mind, we can compute {*c*_*i*1_, …, *c*_*i*7_} from (A.8) to (A.10). For *t >* 7, we can employ (A.7).

- A constraint for 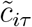 So far, we have not explained how to fix auxiliary daily cases 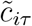. As a starting point, we need to take one constraint into account: As we observe 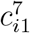, we need to make sure that the sum of 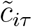 add up to this observed value,

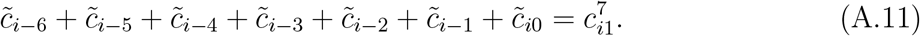

In a second step, we could set auxiliary daily cases to arbitrary numbers, say 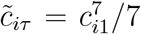 for *τ* = −6 to 0, i.e. we approximate the daily number of cases by one seventh of the first seven-day case rate. This procedure provides us with daily cases {*c*_*i*1_, …, *c*_*i*7_}.

- From case rates to cases - check To be on the safe side, let us now check whether we can recover case rates from our artificial daily cases. Again, the case rate is defined in (A.1), 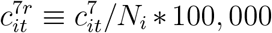. Computing the sum of cases over seven days yields

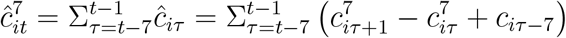

where the second equality employed (A.7). Summing and subtracting yields

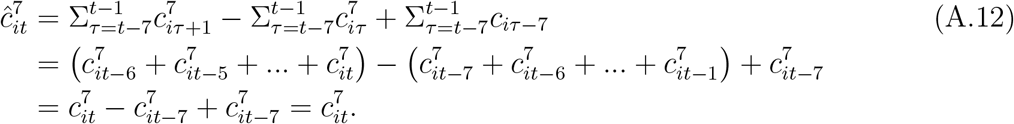

Hence, the computed daily cases allow to recover the original case rates.

An alternative verification approach sums over daily cases in the first week. Employing (A.8) to (A.10), we get

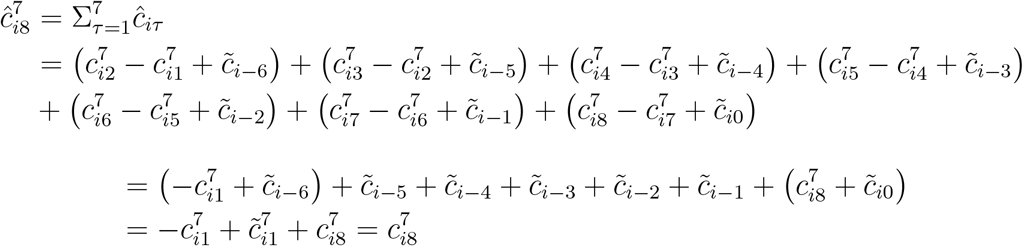

where the latter step assumes that the constraint (A.11) holds,

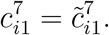

- How to avoid negative daily cases by fixing the unknown cases 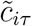 It turned out, however, that the above approach can easily lead to negative cases *c*_*it*_. To understand this, we study this structure in more detail. We find that the level of *c*_*it*_ for *t* ∈ {1, 8, 15, …} is determined by the unknown 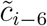. The level of *c*_*it*_ for *t* ∈ {2, 9, 16, …} is determined by the unknown 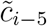 and so on. An overview is in table 5.

We adjust 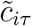, in addition to the constraint (A.11), by computing the minimum of cases by sequences from table 5 and by county *i*. Formally, we need 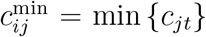 for all counties *i* where *c*_*jt*_ is the number of cases for series *j* or, simply speaking, for weekday *j*. We then adjust the corresponding 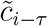 accordingly.

In practice, it turned out that for almost any community, cases were missing. As figure 11 shows, the sum of minima over weekdays is negative for most communities. Hence, this adjustment scheme does not work. There is measurement error.

### A.5 Discussion

#### A.5.1 Percentage deviation

The left panel of figure 12 shows the percentage by which the seven-day case rate is higher (or lower) than in its control region. This left panel can be compared with figure 1 in the main text. The right panel of 12 displays the percentage deviation of cumulative cases from figure 2 in the main text.

#### A.5.2 Cumulative daily cases

Table 6 presents the predictor balance belonging to the second SCM analysis based on cumulative daily cases.

An alternative to figure 2 in the main text is provided by figure 13. It adds a right panel showing confidence intervals. Even though there is a difference in cumulative cases between Augustusburg and its synthetic twin, it is not statistically significant at the 5% level.

#### A.5.3 Robustness ‘leave one out’ analysis

This section complements our robustness analysis in the main part by presenting the figure belonging to the ‘leave one out’ analysis.

